# Single-cell RNA-sequencing reveals predictive features of response to pembrolizumab in Sézary syndrome

**DOI:** 10.1101/2022.03.29.22272994

**Authors:** Tianying Su, Nirasha Ramchurren, Steven P. Fling, Youn H. Kim, Michael S. Khodadoust

## Abstract

The PD-1 inhibitor pembrolizumab is effective in treating Sézary syndrome, a leukemic variant of cutaneous T-cell lymphoma. Our purpose was to investigate the effects of pembrolizumab on healthy and malignant T cells in Sézary syndrome and to discover characteristics that predict pembrolizumab response. Samples were analyzed before and after 3 weeks of pembrolizumab treatment by single-cell RNA-sequencing of 118,961 peripheral blood T cells isolated from six Sézary syndrome patients. T-cell receptor clonotyping, bulk RNA-seq signatures, and whole exome data were integrated to classify malignant T-cells and their underlying subclonal heterogeneity. We found that responses to pembrolizumab were associated with lower *KIR3DL2* expression within Sézary T cells. Pembrolizumab modulated Sézary cell gene expression of T-cell activation associated genes. The CD8 effector populations included clonally expanded populations with a strong cytotoxic profile. Expansions of CD8 terminal effector and CD8 effector memory T-cell populations were observed in responding patients after treatment. We observed intrapatient Sézary cell heterogeneity including subclonal segregation of a coding mutation and copy number variation. Our study reveals differential effects of pembrolizumab in both malignant and healthy T cells. These data support further study of *KIR3DL2* expression and CD8 immune populations as predictive biomarkers of pembrolizumab response in Sézary syndrome.

## Introduction

Immune checkpoint blockade immunotherapy has been shown to produce favorable outcomes in a variety of cancers. Pembrolizumab is a monoclonal antibody against PD-1, an immune checkpoint molecule that induces T-cell exhaustion. Blockade of PD-1 can switch exhausted T-cells to a more invigorated state, in which they can recognize and eliminate cancer cells^1^.

Sézary syndrome is a leukemic subtype of cutaneous T-cell lymphoma typically derived from CD4 central memory T-cells^2^. Pembrolizumab was associated with a 27% overall response rate in patients with Sézary syndrome, and these responses appeared to be especially deep and durable ^3^. Thus, biomarkers of response to pembrolizumab in patients with Sézary syndrome are needed to select the subset of patients who are most likely to experience long-lasting responses. The two current proposed biomarkers of response are not readily translatable because they are either rare and limited to mycosis fungoides such as PD-L1 structural variants ^4^ or require complex spatial analysis from multiplexed imaging data ^5^.

A major obstacle in relating potential biomarkers to clinical outcome is accurately distinguishing the malignant CD4+ T-cells of Sézary syndrome from other similar CD4 cell populations. This is of particular importance in assessing the response to PD-1 inhibition, where the target PD-1 is not only expressed on the immune cells of interest but also by the Sézary cells themselves^6^. Another key challenge in biomarker identification is the underlying tumor heterogeneity of Sézary syndrome. Sequencing of the clonal T-cell receptor (TCR) represents the most specific method to identify the malignant T-cells. Single-cell RNA-sequencing (scRNA-seq) paired with single-cell sequencing of the TCR enables both the assessment of the clonal heterogeneity of Sézary syndrome and the discrimination of malignant from normal T-cells.

In this study, we sought to identify peripheral blood biomarkers of response to pembrolizumab and to decipher the differential effects of pembrolizumab on healthy-T cells versus Sézary cells. We conducted scRNA-seq with paired TCR sequencing on 118,961 T-cells isolated from three Sézary syndrome patients that responded to pembrolizumab treatment and three patients that failed to respond. We analyzed transcriptomic and inferred genomic features, such as mutations and copy number variations, to better understand the immunological, transcriptional, and genomic characteristics that underlie pembrolizumab response.

## Materials and methods

### Human subjects and sample collection

Cryopreserved peripheral blood mononuclear cells were collected as part of a phase II clinical trial of pembrolizumab treatment for mycosis fungoides and Sézary syndrome as previously described (NCT02243579)^3^. For each patient, PBMCs were obtained for before the first dose of pembrolizumab (pre-treatment, Pre), and three weeks later, prior to administration of the second dose of pembrolizumab (Cycle 2, C02). Selected additional samples from later timepoints were analyzed when available. The best clinical response was assessed by consensus global response criteria ^7^ according to the trial protocol. Patients that had either complete or partial response in the clinical trial were classified as “responder” in this study, and patients that had either progressive disease or stable disease were classified as “non-responder”. Written informed consent was obtained from all enrolled patients in accordance with the Declaration of Helsinki. This study was approved by the Institutional Review Board at Stanford University.

### T-cell isolation and scRNA-seq

Cyropreserved PBMCs were thawed through sequential dilution. Live T cells were then immediately isolated using the Human Pan T cell Isolation Kit (Miltenyi Biotec) followed by Dead Cell Removal Kit (Miltenyi Biotec). Cells were resuspended into CryoStor® CS10 freezing medium (STEMCELL Technologies) and transferred into a cold (4°C) Nalgene® Mr. Frosty® Freezing Container (ThermoFisher Scientific) which was immediately placed into − 80°C freezer for four to 24 hours before long-term storage in liquid nitrogen. ScRNA-seq libraries were generated from the cryopreserved T cells using the Chromium Next GEM Single Cell 5’ v2 chemistry (10x Genomics) according to manufacturer’s instructions by MedGenome Inc. (Foster City, CA, USA). TCR sequencing was performed from the barcoded cDNA using the Chromium Single Cell Human TCR Amplification kit following the manufacturer’s instructions by MedGenome Inc. (Foster City, CA, USA). Libraries were sequenced on a NovaSeq 6000 (Illumina), and generation of FASTQ files from BCL file using Cell Ranger (10x Genomics) mkfastq function.

### Alignment and counting of reads

Using the count function in Cell Ranger 4.0.0 (10x Genomics), the gene expression FASTQ files were aligned to the GRCh38 reference transcriptome pre-built by 10x Genomics (version GRCh38 2020-A). Using the vdj function in Cell Ranger 4.0.0, the TCR FASTQ files were aligned to the GRCh38 VDJ reference transcriptome pre-built by 10x Genomics (version GRCh38-alts-ensembl-4.0.0).

### Quality control and clustering of cells

All analysis were performed using Seurat 3.1.4 ^8^ with R 3.6.1 unless otherwise specified. For each dataset, features detected in at least three cells were included in the analysis. Cells that satisfied the following quality control standards were included in the analysis: more than 200 but less than 99.9^th^ percentile for number of features detected, less than 97.5^th^ percentile for percentage of counts belonging to mitochondrial genes, less than 99^th^ percentile for percentage of counts belonging to ribosomal genes, did not express markers of cell types other than T cells. The NormalizeData function was ran to normalize each dataset. The FindVariableFeatures function was used to find differentially expressed genes for clustering. All 15 datasets were then integrated together via the FindIntegrationAnchors and IntegrateData functions. Afterwards, ScaleData function was used to scale the data. RunPCA, RunUMAP, FindNeighbors, and FindClusters functions were performed on the integrated dataset to cluster the cells. In total, 118,961 cells passed all quality control metrics and were included in the analyses.

### Computational identification of T-cell subtypes

Sézary cells were purified via fluorescent-activated cell sorting based on their CD4 and CD26 expression from cryopreserved PBMCs collected prior to pembrolizumab treatment. Total RNA was extracted. RNA-seq libraries were made using SMARTer stranded total RNA-seq Kit v2, pico input kit (Takara) and sequenced on HiSeq 4000 (Illumina). This bulk RNA-seq data was used to build reference transcriptional profiles of Sézary cells for each patient. Bulk RNA-seq data from NCBI SRP125125 ^9^ was used to build reference transcriptional profiles of all other T-cell subtypes. Rsubread 2.0.1 ^10^ was used to align the bulk RNA-seq FASTQ files to the GRCh38 reference transcriptome pre-built by 10x Genomics (version GRCh38 2020-A). To construct a personalized reference transcriptional profile of the T-cell subtypes customized with the transcriptional profile of the Sézary cells from the same patient, a copy of the resulting counts matrix was duplicated for each patient and the counts for the Sézary cells from all other patients were eliminated from the counts matrix. The counts matrices were then normalized using DESeq2 1.26.0 ^11^. Using SummarizedExperiment 1.16.1, each patient’s normalized counts matrix was then made into a “logcounts” transcriptional profile reference for SingleR. For each patient, SingleR 1.0.6^12^ was then used to assign each cell a T-cell subtype identity based on the similarity between the cell’s transcriptional profile and the reference transcriptional profile just built.

### Determination of TRB CDR3 frequencies

Cells with more than one TCR beta (TRB) CDR3 detected or no TRB CDR3 detected were excluded. For each patient, the number of cells with a particular TRB CDR3 were summed for Pre and C02 datasets combined. For each patient, the top clone was defined as the TRB CDR3 with the most number of cells from this analysis and the second clone is the second most common TRB CDR3. For the second top clone, two patients (Pt4 and Pt6) had two second top clones that are equal in number and these were combined together. For all subsequent analyses, the “unknown TRB CDR3” category includes cells with either no TRB CDR3 detected or more than one TRB CDR3 detected.

### Differential gene expression of top clone: non-responder to responder

The FindMarkers function was used to compare top-clone cells from all non-responders to those of all responders. Genes with an adjusted *p*-value (after Bonferroni correction using all features in the dataset) smaller than 0.05 were considered to be significant. To avoid identification of genes driven by one sample or patient, we performed pairwise comparisons using the FindMarkers function between the top-clone cells from each responder and non-responder from pretreatment time points. Genes with an adjusted *p*-value (after Bonferroni correction using all features in the dataset) smaller than 0.05 were considered to be significant. Highlighted genes were differentially expressed in the same direction (upregulated or downregulated) for all non-responder versus responder pairwise comparisons.

### Differential gene expression of top clone: Pre to C02

For each patient, the FindMarkers function was used to compare top-clone cells in Pre to those in C02. Genes with an adjusted *p*-value (after Bonferroni correction using all features in the dataset) smaller than 0.05 were considered to be significant. A statistical overrepresentation test was performed using the PANTHER Pathways annotation data set (PANTHER 16.0 ^13^) for the differentially expressed genes from Pre to C02 for each patient. Fisher’s Exact test was performed, and pathways with a False Discovery Rate of less than 0.05 were considered statistically significant.

From the significant genes, we highlighted genes that met both of the following criteria: 1) found to be differentially expressed in the same direction between Pre to C02 in at least a total of two out of three patients of the same response category, 2) in all three patients of the opposite response category, the gene was either not significantly differentially expressed, or differentially expressed in the opposite direction.

### Cell cycle phase analysis

For each dataset, the CellCycleScoring function was used to calculate for each cell an S phase score and a G2M phase score based on expression of S phase and G2M phase markers (preloaded with the Seurat package) and also to assign each cell a cell cycle phase. The percentage of top clone in G2M was added to those in S phase for each Pre and C02 dataset in each patient. A paired two-tailed *t*-test was used to calculate the *p*-value.

### Identification of second clones’ markers

Cells from Pre and C02 timepoints were combined for each patient and the top-clone cells were excluded. The FindMarkers function was used to find differential gene expression of second-clone cells comparing to cells with other TRB CDR3s for Pre and C02 combined. Genes with an adjusted *p*-value (after Bonferroni correction using all features in the dataset) smaller than 0.05 were considered to be significant and were included for further analyses. From the significant genes, we included only those genes that were significantly changed in the same direction between Pre and C02 for all three responders. Genes that either have cytotoxicity functions or are markers of cytotoxic or effector T cells according to previous studies ^14-18^ were highlighted.

### Calculation of effector scores and cytotoxicity scores

A list of 454 genes with increased expression in CD8 naive cells as compared to CD8 effector memory T cells was used to create an effector score^19^. A list of 16 genes defining a cytotoxicity signature was used to create a cytotoxicity score ^20^. For each dataset, the AddModuleScore function was then used to calculate the effector score and cytotoxicity score for each cell based on its expression of the genes in these lists respectively.

### Quantification of changes in CD8 terminal effector and CD8 effector memory population size

Percentages of non-Sézary cells that were identified as CD8 terminal effector or CD8 effector memory were summed and their change from Pre to C02 was calculated for each patient. A two tailed and heteroscedastic *t*-test was used to calculate the *p*-value.

### Single nucleotide variants analysis

A list of somatic variants were determined from prior exome sequencing for all patients except Pt6^3^. For each patient’s Pre dataset, cb_sniffer^21^ with Python 3.8.5 was used to detect UMIs and cell barcodes with coverage overlying variant genomic positions. Variants were filtered for those with above 10,000 total UMIs detected at the position of the variant. Cells with at least one variant UMI were assigned as containing the mutation. If no variant UMIs were present then they were assigned as wild type for the mutation if there were six or more reference UMIs. Cells with five or fewer reference UMIs and no variant UMIs were categorized as “unknown”.

### Copy number variation analysis

InferCNV 1.7.1 in R 4.0.2 was used to computationally infer the copy number variation for each patient’s top-clone cells from pretreatment time points. The cells with TRB CDR3s other than those of the top or second top clone were used as a reference to establish a baseline upon which copy number variations of the top-clone cells were calculated.

## Results

### scRNA-seq clustering and identification of Sézary cells

To determine the differential effects of pembrolizumab on malignant and healthy T-cells, and to find features that predict response to pembrolizumab, we analyzed peripheral blood T-cells from three patients with response to pembrolizumab and three patients with no response (either progressive disease or stable disease). scRNA-seq and single cell TCR sequencing was performed on a total of 118,961 T-cells isolated from 15 samples collected before and during pembrolizumab treatment (Fig. 1a and Fig. 1b). We analyzed the transcriptome of these cells, as well as inferred various genomic features such as mutations and copy number variations from the scRNA-seq data (Fig. 1a), to gain better understanding of the immunological, genomic, and transcriptomic features that underlie pembrolizumab response.

**Fig 1.**
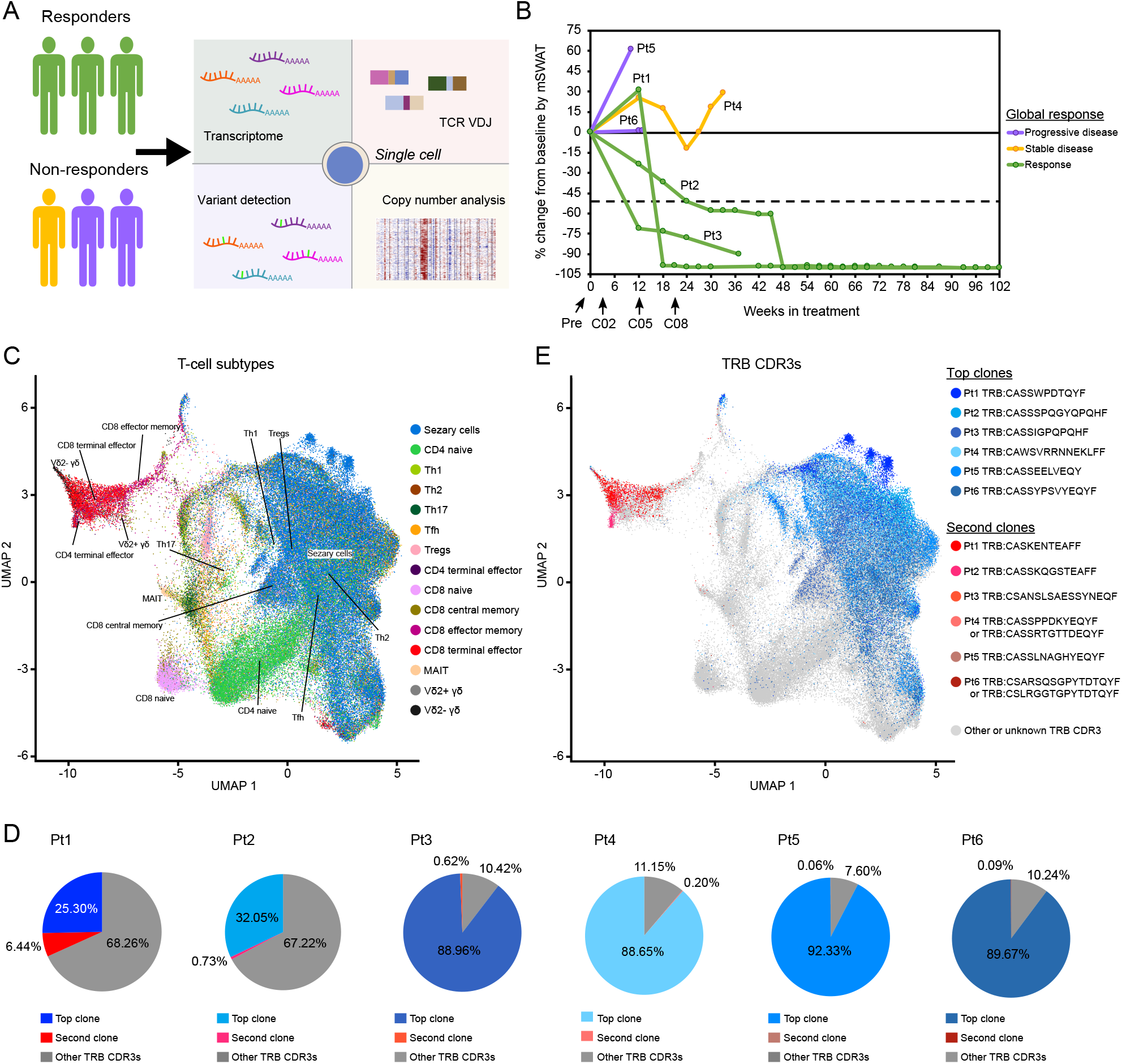
Experiment schematic and clustering of cells. **a** Experiment schematic. **b** Skin response for each patient as measured by modified Severity Weighted Assessment Tool (mSWAT). **c** Uniform Manifold Approximation and Projection (UMAP) of all cells in the analysis, colored by computationally inferred T-cell subtype. **d** Frequency of the two most common TRB clonotypes in each patient inclusive of both Pre and C02 timepoints. **e** UMAP of all cells in the analysis, colored by TRB clonotype. Top clones (in shades of blue) and second most common (in shades of red) for each patient are highlighted in color.

Although TCR clonotyping has traditionally been considered the gold standard to identify malignant T-cells in cutaneous T-cell lymphomas (CTCL), recent observations have suggested that the intratumoral heterogeneity of CTCL may include multiple TCR clonotypes ^22,23^. In addition to a clear dominant TCR clonotype, we noted multiple high frequency TCR clonotypes in several samples as high as 6.4% of all T-cells (Fig. 1d), raising the possibility of multiple Sézary clonotypes. Therefore, to distinguish malignant T-cells from normal T-cells, we incorporated both transcriptomic and TCR clonotypic data to ensure accurate assignment of cell type.

Because of the marked interpatient heterogeneity of Sézary syndrome^24-26^, we used bulk RNA-seq data on immunophenotypically sorted Sézary cells to determine personalized reference transcriptomes for each patient’s Sézary disease and merged these with previously defined references for 29 immune cell types^9^. We then computationally assigned each cell to one of the reference cell types (Fig. 1c). We related the resulting cell classification to the two most frequent TRB CDR3 clonotypes for each patient.

For all patients, cells with the most common TRB CDR3 clonotype (“top clone”) were classified as Sézary cells (Fig. 1e, Fig. 1c). In contrast, cells with the second most common TRB CDR3 clonotype (“second clone”) had a distinct transcriptomic profile. Thus, it appears that the multiple high frequency TCR clones observed in these patients was due to expansion of non-malignant T-cells.

### Sézary gene expression associates with response to pembrolizumab

Because the top clone invariably corresponded with Sézary cells, we used the TRB CDR3 to identify Sézary cells for further analysis. Sézary cell markers were consistent with previous studies (Fig. 2a)^27-29^. We next analyzed pretreatment samples to identify the genes differentially expressed within Sézary cells between responding and non-responding patients (Fig. 2b, Supplementary Table S1). We found three genes consistently differentially expressed between every responding and non-responding patient (Supplementary Table S2). Responses to pembrolizumab were associated with lower expression of *KIR3DL2* and higher expression of *KIF5B* and *HSB17B11* within the Sézary cell population.

**Fig 2.**
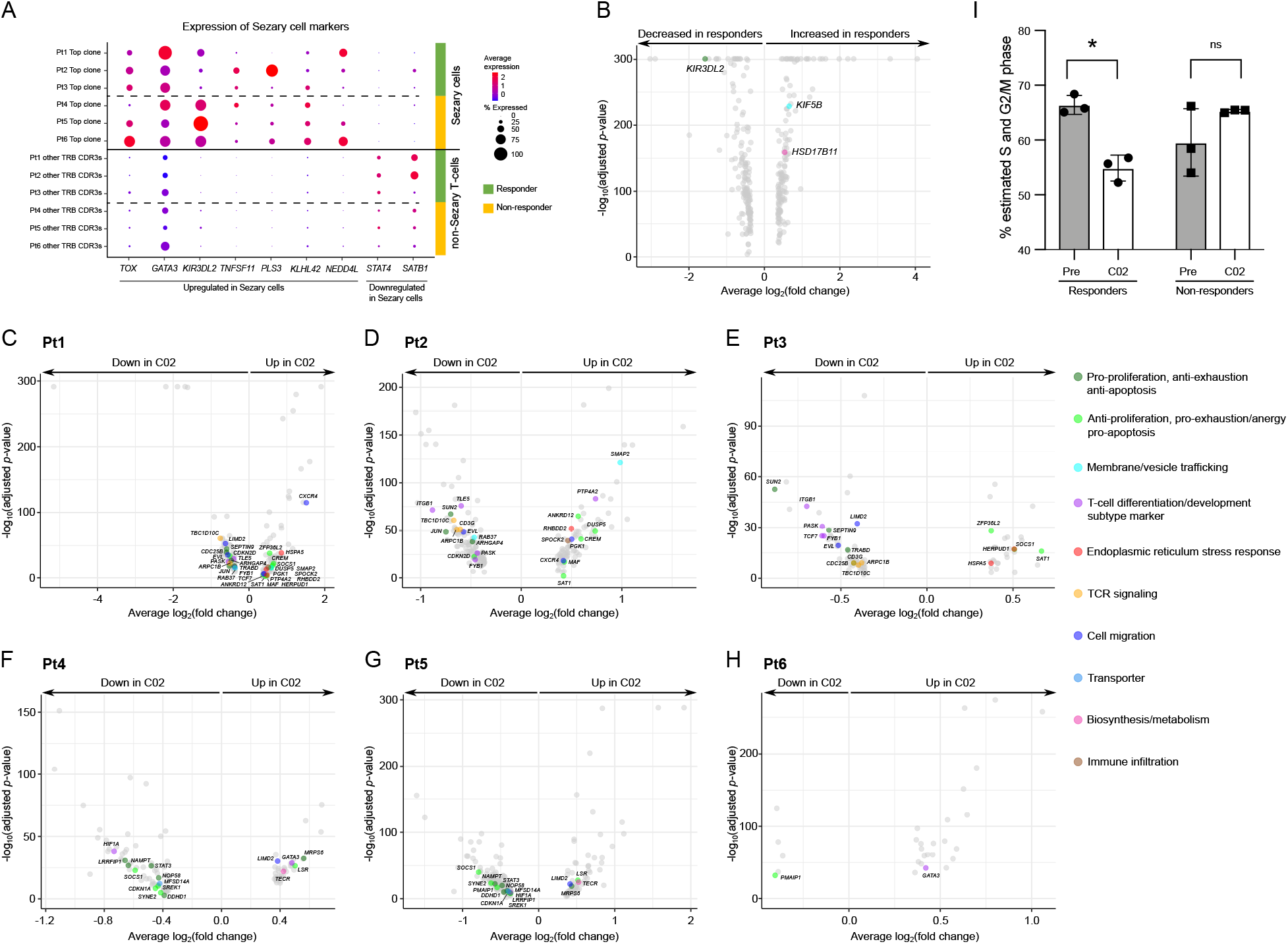
Sézary cell (defined by TCR clonotype) characteristics. **a** Expression of known Sézary cell markers are shown for the Sézary (top) and non-Sézary (bottom) cells. Dot color depicts average expression and dot size depicts the percentage of cells with detectable expression within the cell population **b** Differential gene expression comparing responders and non-responders. Sézary cells from the pretreatment timepoint were analyzed. Genes that were differentially expressed in the same direction in all individual non-responder to responder pairwise comparisons for the Pre datasets are highlighted. **c-h** Differential gene expression within the Sézary cell population comparing Pre to C02. Genes that were significantly differentially expressed in the same direction in either multiple responders or non-responders are highlighted. **i** Percentage of Sézary cells that were computationally determined to be in either the G2M or S phase of the cell cycle. * *p* < 0.05; ns, not significant.

### Effects of pembrolizumab treatment on Sézary cells

Because these Sézary cells are known to have varying expression of PD-1^3^, we sought to determine the effects of pembrolizumab treatment on the Sézary cell population. We compared samples obtained prior to treatment and to those collected three weeks after the first dose of pembrolizumab.

Pembrolizumab treatment modulated expression of genes associated with T-cell activation and apoptosis signaling within the Sézary cell population (Fig. 2c-h, Supplementary Table S3-S4). Pembrolizumab significantly increased *SOCS1* expression in the Sézary cells of two responding patients, but significantly decreased *SOCS1* expression in two non-responding patients. In all responding patients, pembrolizumab treatment was associated with a decrease in expression of the actin cytoskeletal genes *FYB1, TBC1D10C*, and *ARPC1B*. Additionally, in responders, we observed decreased expression of *TCF7* by Sézary cells after pembrolizumab, which has been shown to be required for self-renewal of both CD8 and CD4 T-cells ^30,31^.

Treatment of T-cell malignancies with immune checkpoint inhibitors presents a theoretic risk of disease hyperprogression through release of T-cell inhibition. Examples of hyperprogression with PD-1 treatment have been observed in adult T-cell leukemia and peripheral T-cell lymphomas ^32,33^ and stimulation of leukemic CTCL with nivolumab increases malignant T-cell proliferation *ex vivo* ^34^. To further evaluate the effect of pembrolizumab on Sézary cell proliferation, alterations in the cell cycle after three weeks of treatment were computationally inferred for Sézary cells. The percentage of Sézary cells in the more proliferative phases of the cell cycle (G2M and S) decreased significantly for responders while no significant change was observed for the non-responders (Fig. 2i).

### Effects of pembrolizumab treatment on expanded cytotoxic clonotypic cells

We next explored the phenotype of the high frequency, non-malignant T-cell clones by analyzing the second most common T-cell clonotype in each patient. Differential gene expression comparison to other T-cell clonotypes revealed that the expanded second clones were associated with markers of cytotoxic lineage (Fig. 3a, Supplementary Table S5). Together the clustering of the second clones with CD8 terminal effector T cells on the UMAP projection (Fig. 1e, Fig. 1c), these results indicated that the second-clone cells were most likely cytotoxic CD8 terminal effector T cells.

**Fig 3.**
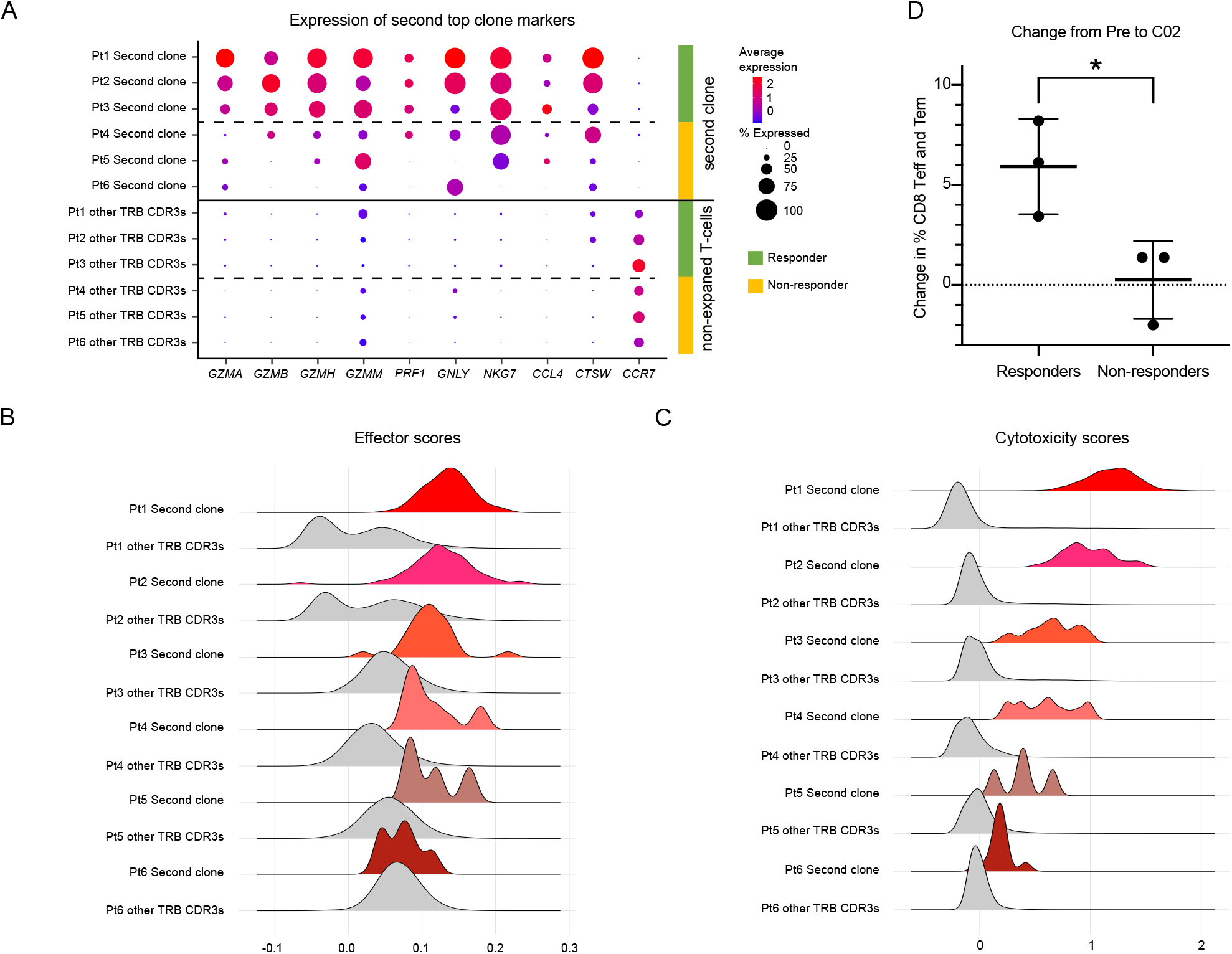
Characterization of expanded clones (cells with the second most common TRB CDR3) from each patient. **a** Expression of cytotoxicity and effector T-cell are shown for the second clone (top) and all other non-Sézary T-cells (bottom). **b** Effector scores of the second clone and cells with other TRB CDR3s in each patient. **c** Cytotoxicity scores of the second clone and cells with other TRB CDR3s in each patient. For **a-c** analyses were performed on cells from both Pre and C02 timepoints combined. **d** Change from Pre to C02 in CD8 terminal effector or CD8 effector memory T cells as a percentage of non-Sézary T-cells in responders and non-responders. *: *p* < 0.05, Teff: terminal effector T cell, Tem: effector memory T cell.

We noted increased expression of cytotoxicity markers in the second clones of responding patients as compared to non-responders (Fig. 3a). To further investigate the effector and cytotoxicity characteristics of the second clone, an effector score and a cytotoxicity score were calculated for each cell based on previously defined gene signatures ^19,20^. For all patients, the second clone appeared to have higher effector and cytotoxicity scores than cells with other TRB CDR3s (Fig. 3b, Fig. 3c). Both the cytotoxicity and effector score of the second clones appeared to higher in responding patients as compared to those in non-responding patients (Fig. 3b, Fig. 3c).

Proliferation of CD8 T-cells has been associated with response to PD-1 inhibition in other malignancies^35^. Therefore, we tested whether pembrolizumab differentially affected the expansion of CD8 populations in responders versus non-responders. We compared the change in frequencies for computationally inferred T-cell subtypes for each patient after 3 weeks of pembrolizumab treatment. Pembrolizumab treatment yielded significantly greater expansion of the CD8 effector population (terminal effector and effector memory) in responders as compared non-responders (Fig. 3d).

### Intrapatient Sézary cell heterogeneity through integrated single cell analysis of single nucleotide variant and copy number alterations

Sézary syndrome has been shown to have variable genomic intrapatient heterogeneity ^23,36^ typically with multiple identified subclones. Because intratumoral genomic heterogeneity has been correlated with clinical response to immunotherapy in other malignancies ^37^, we leveraged this dataset to explore clonal heterogeneity at a single cell level through integration of TCR clonotype, subclonal copy number alterations and single nucleotide variants.

We searched within our scRNA-seq data for patient-specific single nucleotide variants (SNV) determined through bulk exome sequencing^3^. The detection of these variants by scRNA-seq is strongly dependent on both gene expression and the cDNA position of the variant. Therefore, our analysis was restricted to five variants associated with three patients (Pts 1, 2, and 5. For all mutations, cells expressing the variant were almost exclusively limited to the Sézary TRB clonotype (Fig. 4a-d). Sézary cells displayed heterogeneity for some of the variants (Fig. 4d). For the responding Pt2, the Sézary cells were more clonal with respect to the analyzed variants comparing to the non-responding Pt5 (Fig. 4e).

**Fig 4.**
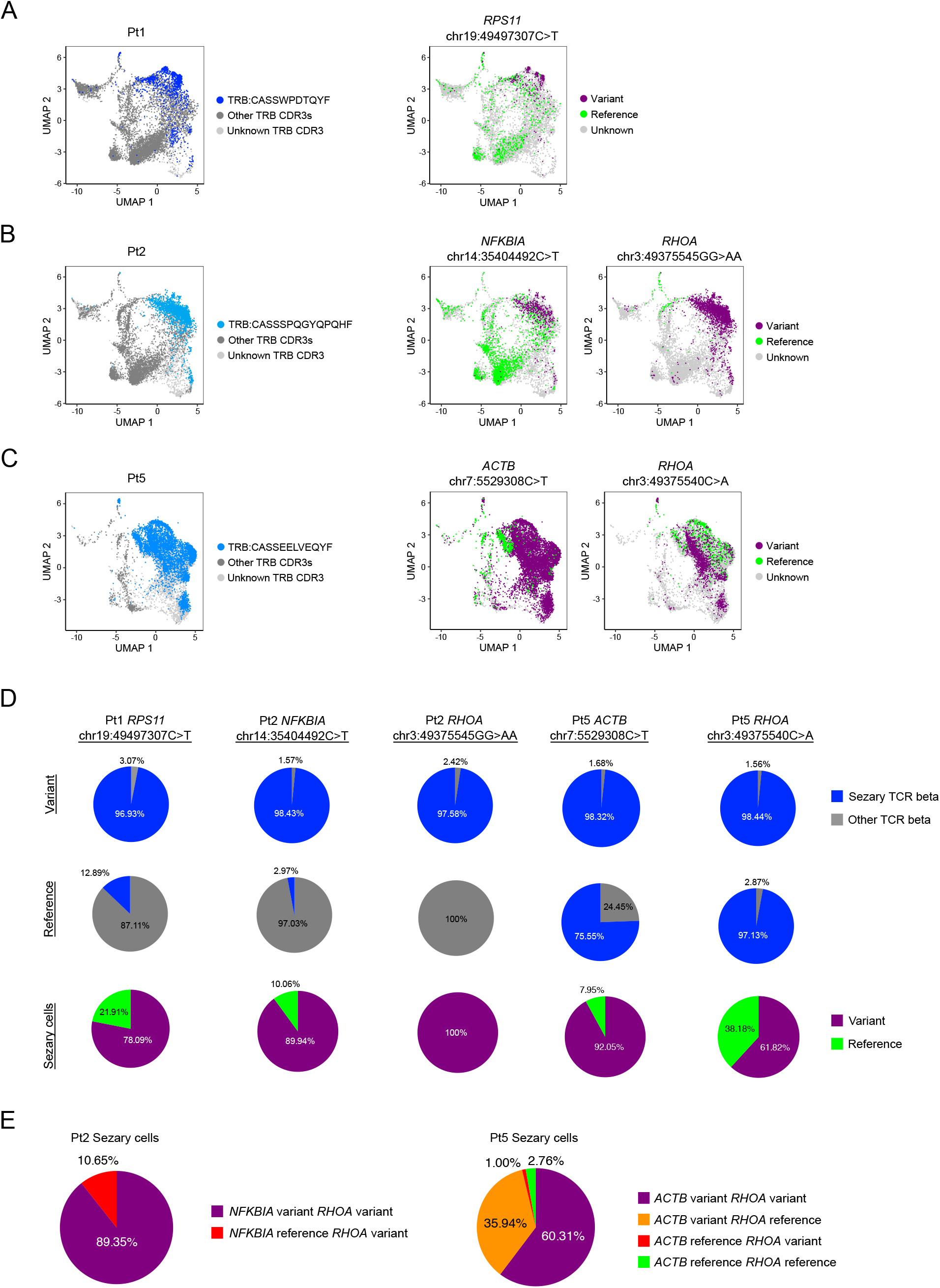
Heterogeneity of Sézary population through detection of single nucleotide variants. **a-c** UMAP visualization highlighting the Sézary clonotype for three patients (left) and patient-specific somatic variants (right). Cells are labeled as containing the variant if there was at least one variant read detected. Cells with six or more non-variant reads and no variant reads are labeled as reference. **d** For each of the mutations shown in **a-c**, The percentage of variant or reference cells identified as Sézary or non-Sézary by TRB clonotype is shown (top and middle rows). The percentage of Sézary cells expressing either the variant or reference sequence is also shown (bottom row). **e** For the two patients with multiple somatic variants detected, the percentage of Sézary cells with one or both variants is shown.

Next, we explored clonal heterogeneity through inferred copy number variants (CNVs) within the Sézary cell populations. Clonal CNVs were present in all patients, including clonal aberrations in chr17 (5 of 6 patients, Fig. 5b-f), which is one of the most common copy number alteration in Sézary syndrome ^38,39^. Additionally, at least one subclonal CNV was identified in each patient. These subclones typically comprised only a small fraction of the total Sézary population. No remarkable difference in copy number variation was observed between responders and non-responders (Fig. 5a-f).

**Fig 5.**
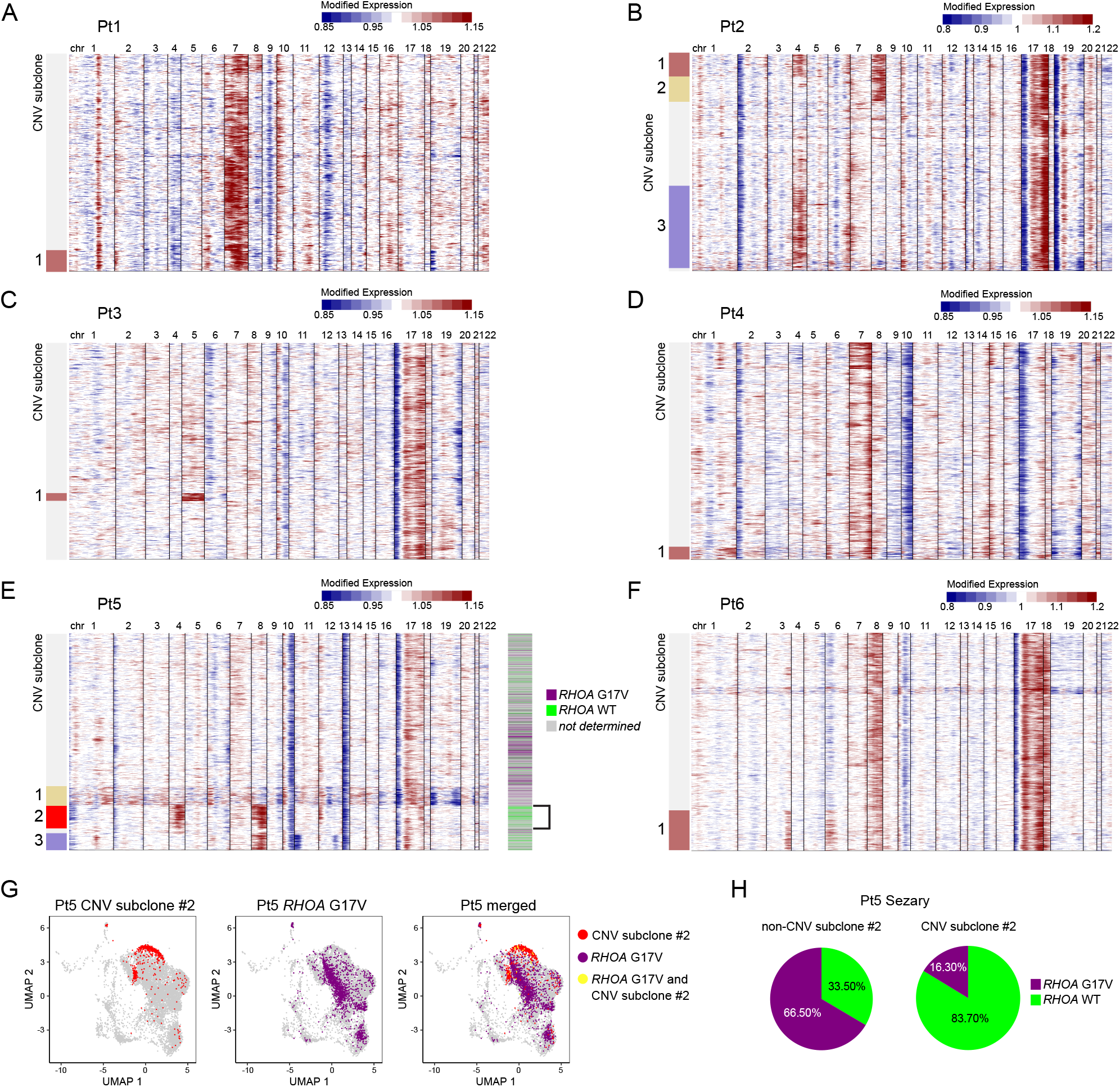
Heterogeneity of the Sézary population through copy number analysis. **a-f** Computationally inferred copy number variation heatmap of the Sézary clones prior to treatment for each patient with each row representing a Sézary cell. Subclones indicate subsets of cells with unique copy number variations. In panel e (right) the genotype of the RHOA^G17V^ for each cell is shown with the bracket corresponding to cells belonging to CNV subclone 2. **g** UMAP plots of the pretreatment sample for Pt5 highlighting cells belonging to CNV subclone 2 (left), expressing the RHOA^G17V^ variant (middle) and merging both characteristics (right). **i** Percentage of RHOA^G17V^ variant in Pt5 Sézary cells with or without the CNV of subclone 2. CNV: copy number variation, WT: wild-type.

To further elucidate the clonal architecture, we integrated cell-specific single nucleotide variants with the inferred copy number variants. We focused on Pt5 where the Sézary cells demonstrated a subclonal RHOA^G17V^ mutation as well as subclonal CNV variants. We found that copy gains in chr4 and chr8 and the RHOA^G17V^ variant appeared to be mutually exclusive and present in independent subclones (Fig 5g-h).

Finally, we explored whether resistance to pembrolizumab was due to selection of specific resistant subclones. However, we found no significant change in the subclonal composition of Sézary cells when comparing pretreatment samples with those collected after three weeks of pembrolizumab treatment (data not shown).

## Discussion

Distinguishing malignant from non-malignant T-cells in CTCL poses a major challenge in identifying potential biomarkers of response to immunotherapy. High dimensional analysis such as mass cytometry or highly multiplexed imaging can more reliably identify malignant T-cells, but still may be imperfect in resolving CTCL cells from similar CD4 T-cell subsets. We used single cell analysis with TCR sequencing to better resolve tumor-intrinsic and tumor-extrinsic features. Three Sézary cell-instrinsic markers were discovered through this approach. High expression of *HSD17B11* and *KIF5B* was associated with responding patient. The short-chain alcohol dehydrogenase gene *HSD17B11* has previously been identified as a serologic tumor antigen in CTCL with 4 out of 6 patients demonstrating serological reactivity against the antigen ^40^. *KIR3DL2* has been consistently been found to be overexpressed in Sézary cells^41^. *KIR3DL2*, according to previous studies, helps Sézary cells avoid activation-induced cell death ^42,43^. Expression of *KIR3DL2* and *PD-1* has been proposed to define three subclasses of Sézary syndrome with distinct skin immune microenvironments including higher numbers of reactive CD8+ T-cells in the setting of low *KIR3DL2* expression ^44^.

We found that pembrolizumab altered expression of TCR signaling pathway genes by Sezary cells. *Ex vivo* studies have suggested *PD-1* inhibition may induce Sézary cell proliferation ^34^. However, our analysis suggests a variable response to *PD-1* blockade. Additionally, expression of genes involved in T-cell differentiation such as *GATA3* and *TCF7* were also significantly changed after treatment with pembrolizumab. *TCF7* has been associated with T-cell “stemness” ^45^. Therefore, it is possible that by decreasing *TCF7* expression, pembrolizumab may inhibit the ability of Sezary cells to continue their renewal in responding patients. The changes in the cell cycle distribution and *TCF7* expression of Sézary cells after pembrolizumab treatment suggests that the response to therapy may at least in part be due to intrinsic effects of pembrolizumab on Sézary cells.

Our data suggest that effector CD8 populations also play a role in mediating the response to pembrolizumab. While tumor infiltrating CD8+ T-cells have consistently been implicated in response to immune checkpoint therapies in solid malignancies ^46^, no correlation was seen between cutaneous CD8 T-cells and response to pembrolizumab in this trial ^3,5^. However, in analyzing circulating lymphocytes, we found peripheral blood CD8 terminal effector and CD8 effector memory T-cell populations expanded in responding patients upon pembrolizumab treatment. Furthermore, we found a marked clonal expansion of CD8 terminal effector cells in patients with Sézary syndrome, reaching a frequency of >6% of all peripheral blood T-cells in one patient with a long-term response to pembrolizumab. This population shared a strong cytotoxic profile that mirrors those seen in clonally expanded peripheral blood CD8 populations in metastatic melanoma^18^. The clonal populations were largest in the three responding patients in our cohort and clonal size of expanded CD8+ T-cell clones has been shown to predict response to immune checkpoint blockade in metastatic melanoma ^18,47^.

Finally, by integrating prior genomic data, we were able to investigate the clonal architecture of these Sézary patients at a single cell level. Recent studies have suggested that CTCL may generate multiple malignant TCR clonotypes. By relating transcriptomes, TCR sequence, copy number variants and single nucleotide variants at the single cell level, we found no evidence of multiple malignant TCR clonotypes. Our data nevertheless did confirm the marked intratumor heterogeneity that has been observed in multiple complementary studies ^23,24,26,36,48^. We further demonstrate the first example of branched evolution in CTCL involving subclonal segregration of a single nucleotide mutation (RHOA G17V) and a copy number variant (gain of chr 4 and 8). We found no significant changes in the subclonal composition in these patients after pembrolizumab treatment. It is likely that one cycle of treatment is too short of a period to detect changes in the clonal heterogeneity.

In conclusion, our study show that responses to pembrolizumab in Sézary syndrome are associated with tumor-intrinsic features and with characteristics of circulating CD8 T cell populations. Expression of the CTCL marker *KIR3DL2* deserves particular consideration as a potential biomarker of response in future studies of immune checkpoint therapy in CTCL. Since low expression of *KIR3DL2* was associated with response, these results raise the possibility that treatment with anti-KIR3DL2 treatment such as lacutamuab ^49^ could synergize with pembrolizumab therapy.

## Data Availability

Data will be made available upon publication of this study in a peer-reviewed journal.

## Author contributions

T.S. and M.S.K. conceptualized this study. N.R., S.P.F., Y.H.K., and M.S.K. performed sample collection and annotation of clinical trial samples. T.S. performed T cell isolation, processing of data, and computational analysis with input from M.S.K. T.S. and M.S.K. performed bulk RNA sequencing. T.S. and M.S.K. wrote the manuscript with all authors providing input.

## Acknowledgements

This work was supported by the NIH under Grant K08 CA207882; NCI under Grant 5UM1CA154967, and the Haas Family Foundation.

## Ethical approval and informed consent

This study was approved by the Stanford University Institutional Review Board (IRB). A written informed consent was obtained from all patients who participated in this study in accordance with the Declaration of Helsinki.

## Supplementary table legends

**Supplementary Table S1**. Details (log*e* fold change, adjusted *p*-value) of the differential gene expression analysis shown in Fig. 2b. Genes that were differentially expressed in the same direction in all individual non-responder to responder pairwise comparisons for the Pre datasets are colored by their gene function. avg loge(FC): natural log of fold change of average expression, p val adj: adjusted *p*-value (after Bonferroni correction using all features in the dataset).

**Supplementary Table S2**. Patient level details of the genes from Fig. 2b and Supplementary Table S1 that were significantly differentially expressed in the same direction in all individual non-responder to responder pairwise comparisons. This table shows the log*e* fold change and adjusted *p*-value for these genes from all those aforementioned individual non-responder to responder pairwise comparisons. A brief description of gene function is shown for each gene. avg loge(FC): natural log of fold change of average expression, p val adj: adjusted *p*-value (after Bonferroni correction using all features in the dataset).

**Supplementary Table S3**. Details (log*e* fold change, adjusted *p*-value) of the differential gene expression analysis shown in Fig. 2c-h. Genes that met these criteria were colored by their gene function: 1) found to be differentially expressed in the same direction between Pre to C02 in at least a total of two out of three patients of the same response category, 2) in all three patients of the opposite response category, the gene was either not significantly differentially expressed, or differentially expressed in the opposite direction. The overrepresented PANTHER pathways in these differentially expressed genes for each patient is shown in the “PANTHER pathways” tab. avg loge(FC): natural log of fold change of average expression, p val adj: adjusted *p*-value (after Bonferroni correction using all features in the dataset), NA: gene was not differentially expressed for the comparison shown, FDR: False Discovery Rate.

**Supplementary Table S4**. Patient level details of the genes from Fig. 2c-h and Supplementary Table S3 that were significantly differentially expressed in the same direction in multiple responders (Responders tab) or non-responders (Non-Responders tab). Each of the highlighted genes satisfied these three criteria: 1) differentially expressed in the same direction between Pre to C02 in at least a total of two out of three patients of the same response category, 2) in all 3 patients of the opposite response category, the gene was either not significantly differentially expressed, or differentially expressed in the opposite direction. This table shows for each gene the log*e* fold change and adjusted *p*-value from each patient’s Sezary cells comparing differential gene expression from Pre to C02. A brief description of gene function is also shown for each gene. avg loge(FC): natural log of fold change of average expression, p val adj: adjusted *p*-value (after Bonferroni correction using all features in the dataset), NA: gene was not differentially expressed for the comparison shown.

**Supplementary Table S5**. Genes that were significantly differentially expressed (adjusted *p*-value smaller than 0.05 after Bonferroni correction using all features in the dataset) in the same direction for all responders, from other cells to the second clone, combining Pre and C02 samples. The log*e* fold change and adjusted *p*-value from each patient’s differential gene expression analysis is shown for each gene. Genes that have cytotoxicity functions or are markers of cytotoxic or effector T cells are highlighted in yellow. avg loge(FC): natural log of fold change of average expression, p val adj: adjusted *p*-value (after Bonferroni correction using all features in the dataset), NA: gene was not differentially expressed for the comparison shown.

